# Windows into leishmaniasis epidemiology in the United States: September 2021 through August 2022

**DOI:** 10.1101/2023.05.23.23290303

**Authors:** Thao T. Truong, Karissa Crawford, Ichih Wang-McGuire, Kendal Jensen, Aisha Mushtaq, Nicole A.P. Lieberman, Frederick S. Buckner, Wesley C. Van Voorhis, Brad T. Cookson, Stephen J. Salipante, Joshua A. Lieberman

## Abstract

Leishmaniasis is a rare disease in the United States, with an estimated annual incidence of dozens of cases occurring primarily in travelers, migrants, and military personnel. True disease incidence is unknown, since leishmaniasis is not a nationally notifiable condition. Here, we describe the results of molecular leishmaniasis over a 1-year interval (September 2021 to August 2022) when our laboratory served as the primary national reference laboratory for molecular diagnosis of civilian leishmaniasis. We tested 218 specimens submitted from 36 states yielding 94/186 (50.5%) positive cases with species or species complex-level identification and 18 novel mini-exon alleles. Most species belonged to subgenus *Viannia* (75.6%) and associated with cutaneous or mucocutaneous disease. Cases were associated with recent travel (18.1%), travel timing unspecified (7.4%), migration (7.4%), remote travel (2.1%), military (1.1%), or unknown history (63.8%). These data illustrate the clinical utility of molecular testing for leishmaniasis and provide unique insight into disease epidemiology.

## Introduction

*Leishmania* parasites are transmitted to humans by bites from infected phlebotomine sandflies. Over 20 *Leishmania* species are described and cause a wide spectrum of clinical disease in humans. Although leishmaniasis is considered a neglected tropical disease, it is endemic to 98 countries and its worldwide distribution is impacted by human conflict, travel, immigration, and displacement (1). *Leishmania* species endemic to the Eastern hemisphere have been geographically categorized as “Old World” *Leishmania* (OWL) whereas those endemic to the Western hemisphere have been termed “New World” *Leishmania* (NWL) (2). Most human infection is caused by species from one of two subgenera. The New World subgenus *Viannia* includes *L. braziliensis* spp. complex and *L. guyanensis* spp. complex, which cause cutaneous and/or mucocutaneous lesions. Subgenus *Leishmania* includes Old World *L. major*, Old World *L. tropica* spp. complex, and New World *L. mexicana* spp. complex, which all cause cutaneous leishmaniasis (CL). The *Leishmania* subgenus additionally includes *L. donovani* (OWL) and *L. infantum* (OWL)*/L. chagasi* (NWL), which are associated with organotropic visceral leishmaniasis (VL), the most fatal clinical manifestation.

Diagnostic testing methods include histopathology to visualize intracellular amastigote stage parasites, *in vitro* culturing from lesions, serology, and molecular detection of parasite DNA (3). However, molecular methods are considered most sensitive, and are often most rapid (2–5). The University of Washington (UW) reference laboratory developed and validated a polymerase chain reaction (PCR) and Sanger sequencing assay to detect and identify *Leishmania* from tissue and body fluid specimens. The approach interrogates the organism’s multicopy mini-exon gene using primers targeting regions conserved across *Leishmania* species (6). The amplified region is sufficiently diverse to allow taxonomic determination of species or species complex (6,7). Along with patient immune status, type and severity of clinical disease, and travel history, knowledge of the infecting *Leishmania* species helps determine the risk for mucocutaneous leishmaniasis (MCL), which can impact treatment and management (3).

*Leishmania* has been considered endemic to the United States since 2015, after several reports of autochthonous CL (8–10). Previous studies of *Leishmania* infection epidemiology in the U.S. have been limited in scope and often lack species-level identification, comprising surveillance of armed forces members, case reports, and multicenter observational studies (11–13). During September 2021 – July 2022, culture, serology, and molecular testing and identification of *Leishmania* species was temporarily discontinued at the U.S. Centers for Disease Control and Prevention (CDC) as part of an agency-wide pause on clinical testing (14), leaving the UW diagnostic reference laboratory the only service providing molecular *Leishmania* testing to the general U.S. population during this period (3). This situation offers a unique and relatively unbiased opportunity to examine molecular testing results for patients with suspected leishmaniasis within the United States. Here, we present the largest survey of *Leishmania* testing in the U.S. performed to date, encompassing 94 positive patient cases with species-level identification from testing specimens from 186 patients submitted over a 1-year period. Our work lends significant insights into descriptive and molecular epidemiology of leishmaniasis in the U.S.

## Materials and Methods

### Assay Validation

We validated a PCR and Sanger sequencing assay targeting the *Leishmania* mini-exon gene (6) under the CLIA ‘88 regulatory framework and guidelines established by the Clinical Laboratory Standards Institute. Assay performance was assessed using 7 cultured isolates (axenic promastigote culture) and genomic DNA purchased from the American Type Culture Collection (ATCC 50129); 13 residual clinical specimens positive for *Leishmania* spp. by broad-range fungal PCR performed in our laboratory and/or by culture and molecular analysis at the CDC Parasitic Disease Branch; and 22 residual clinical specimens negative for *Leishmania* spp. but positive for other pathogens (Table S1). DNA extraction, sequence analysis, and case review were performed as previously described (15,16). Specimens were tested in technical duplicate with positive, negative, and inhibition control reactions. Assay performance was fully reproducible across 3 operators.

### Patient Population

The study cohort included all patients with leishmania testing performed at the UW Molecular Microbiology clinical diagnostic laboratory between the assay’s initial offering on 9/1/2021 and 8/31/2022. The CDC referred leishmaniasis testing to the University of Washington from 10/2021 to 7/2022. Acceptable specimen types included fresh and formalin-fixed paraffin-embedded (FFPE) tissue, body fluids, buffy coat preparations, and (with laboratory director approval) stain-positive peripheral blood.

### Demographic and Clinical Information

Demographic data including sex, age, state from which specimen was submitted, dates of collection/submission, and specimen description/anatomic site were obtained from the laboratory information system. When available, clinical history, histopathology findings, and travel history were obtained from medical records including physician clinicopathologic consultations.

### Ethical Approval

The study was approved by the UW IRB (STUDY00013877).

### Data Analysis

Data were analyzed in R (version 4.2.1) (17). Maps and figures were generated using usmap, tidyverse, cowplot, and ggtree packages (18–21). “Cases” were defined as unique patients, regardless of the number of specimens tested. Chi-square tests were used to compare categorical variables using GraphPad QuickCalcs (22).

Assembled mini-exon sequences were MAFFT aligned in Unipro Ugene v44.0 (23) with published reference sequences representing human-infecting species complexes and curated manually to consolidate mini-exon alleles exhibiting minor length variation. Trees were generated by IQ-Tree with automatic substitution model optimization (24) and visualized with ggtree (25).

### Data availability

Sequences have been deposited to Genbank (accessions OR026045-OR026086) with metadata available in BioSample accessions SAMN35159702 - SAMN35159803 (BioProject PRJNA974035). Deidentified, patient-specific molecular data are available upon request with appropriate human subject approvals.

## Results

### Assay Validation

We determined assay performance for a PCR and Sanger sequencing assay targeting the *Leishmania* mini-exon gene (6) using extracted DNA from axenic promastigote culture and residual clinical specimens known to be positive for *Leishmania* spp. or common and/or morphologically similar pathogens (Table S1). The assay’s 95% limit of detection (LOD) was established as 1 genome per reaction by testing serial dilutions of purified genomic DNA into testing matrix (Table S2). Sensitivity of testing for DNA extracted from cultured promastigote cell pellets, purified parasite DNA, and patient specimens was 90.5%. Species complex identification was fully concordant with CDC gold-standard testing. Specificity was 100%, with no cross-reactivity with human DNA or other tested pathogens (Table S1).

### Testing cohort

We clinically tested 218 specimens from 186 patients over the one-year period from September 2021 through August 2022. 94 patients (50.5%) had at least 1 specimen positive for *Leishmania* (Table 1). Specimens were submitted from 36 U.S. states and the territory of Puerto Rico, as well as Quebec, Canada and the Dominican Republic (Figure 1). California, Florida, and Texas submitted the most cases for testing, with corresponding positivity rates of 48.5% (16/33), 50.0% (8/16), and 41.2% (7/17) (Figure 1D). Overall case and specimen positivity rates were 50.5% and 47.7% over the study period (Figure 2A, Table 1, Table 2). The months with the highest case volumes were June, May, and February 2022 (Figure S1). Median test turnaround time was 3.19 days (IQR 1.32 - 3.91) from specimen receipt.

**Figure 1.**
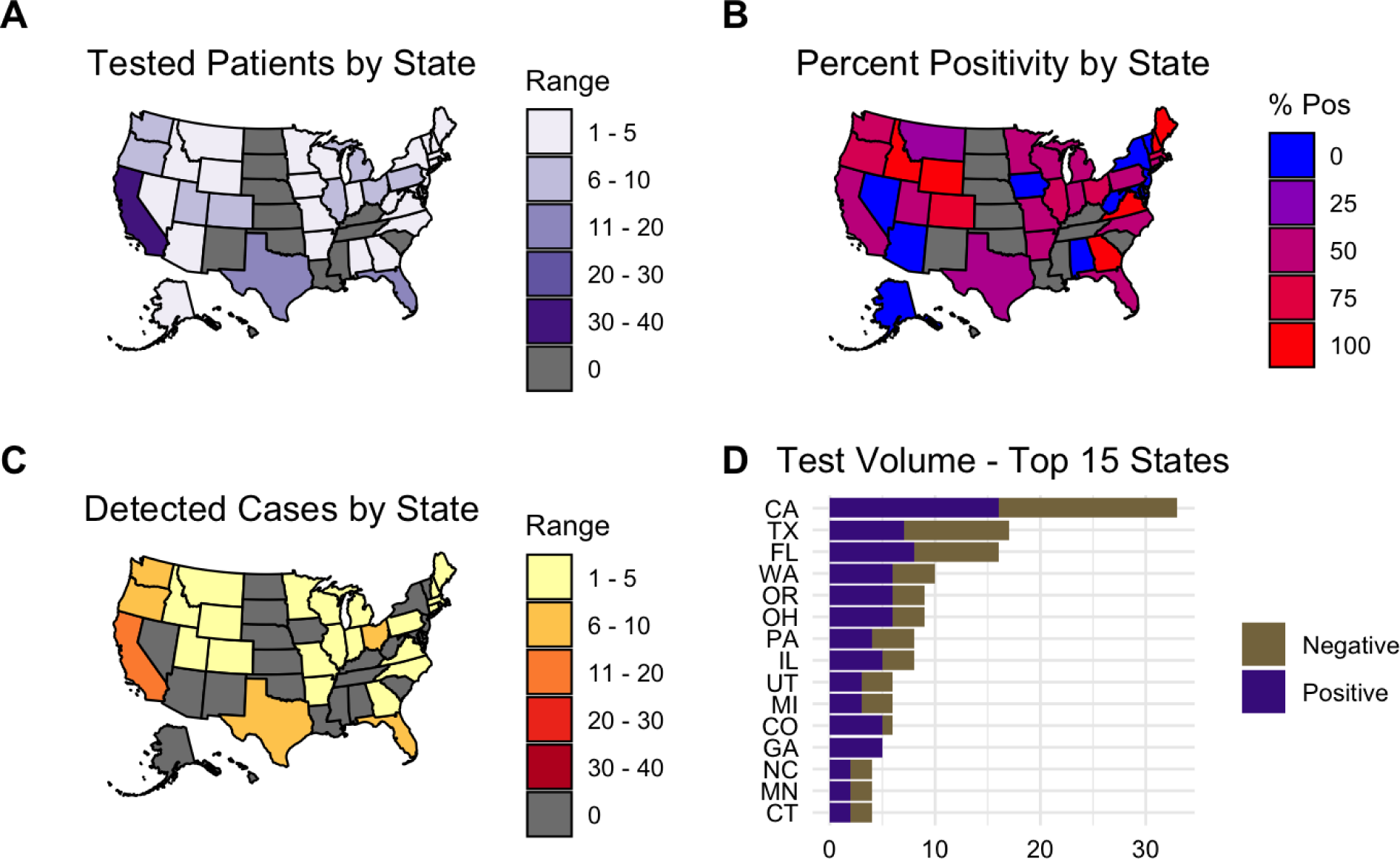
Geographic distribution of patients whose specimens were submitted to our laboratory for *Leishmania* testing from September 2021 to August 2022. Not depicted in this figure: 1 case from Quebec, Canada, 1 case from Puerto Rico. 1 case from the Dominican Republic was submitted by an institution in Florida. (A) The number of patients tested by each state in the U.S. (B) The positivity rate per state during this time period. States shaded in grey did not submit any specimens. (C) The number of patients who tested positive for *Leishmania* by each state. (D) List of the top 15 states by total test volume.

**Figure 2.**
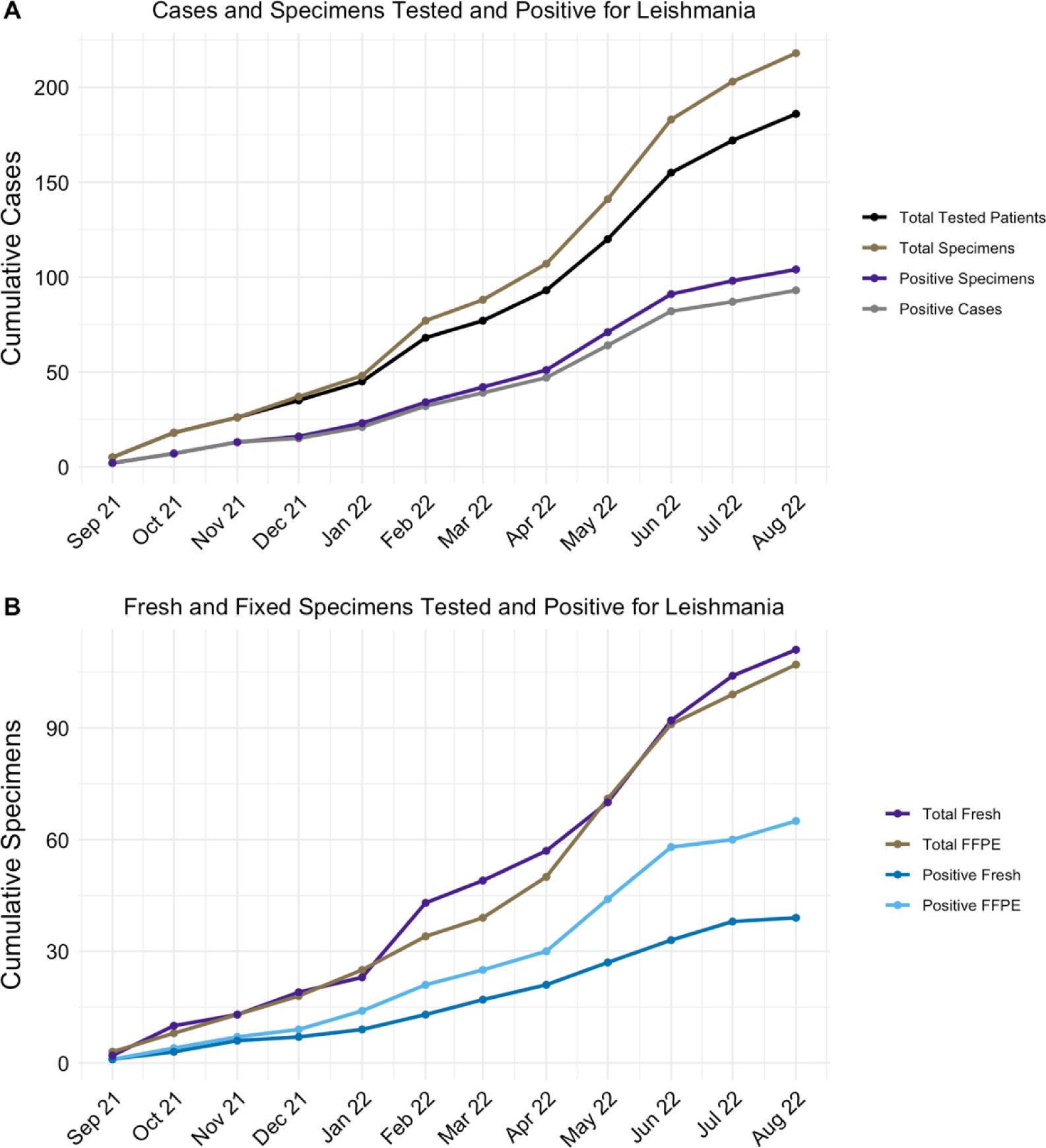
Cumulative number of specimens and cases from September 2021 to August 2022. Cumulative testing showing (A) total and positive volume of patients and specimens during each month and (B) total and positive volume of fresh vs formalin-fixed paraffin-embedded specimens during each month.

**Table 1.** Demographic characteristics of patients with positive *Leishmania* PCR results.

| **Characteristic** | **# cases tested** | **# cases positive** | **(%) positive by characteristic** | **(%) positive out of total positive cases (n = 94)** |
| --- | --- | --- | --- | --- |
| **Total tested** | 186 | 94 | (50.5) | - |
| **Age group** |  |  |  |  |
| < 20 | 30 | 19 | (63.3) | (20.2) |
| < 10 | 15 | 11 | (73.3) | (11.7) |
| 20-39 | 60 | 34 | (56.7) | (36.2) |
| 40-59 | 57 | 25 | (43.9) | (26.6) |
| 60-79 | 35 | 15 | (42.9) | (16.0) |
| > 80 | 4 | 1 | (25.0) | (1.1) |
| **Sex** |  |  |  |  |
| Male | 107 | 60 | (56.1) | (63.8) |
| Female | 76 | 32 | (42.1) | (34.0) |
| Unknown | 3 | 2 | (66.7) | (2.1) |
| **Travel history** |  |  |  |  |
| Travel history |  |  |  |  |
| Migration | 10 | 7 | (70.0) | (7.4) |
| Travel ≤6 months | 21 | 17 | (81.0) | (18.1) |
| Travel >6 months | 4 | 2 | (50.0) | (2.1) |
| Travel, timing not specified | 13 | 7 | (53.8) | (7.4) |
| Military | 1 | 1 | (100.0) | (1.1) |
| No travel history | 1 | 0 | (0) | (0) |
| Unknown | 136 | 60 | (44.1) | (63.8) |

**Table 2.**
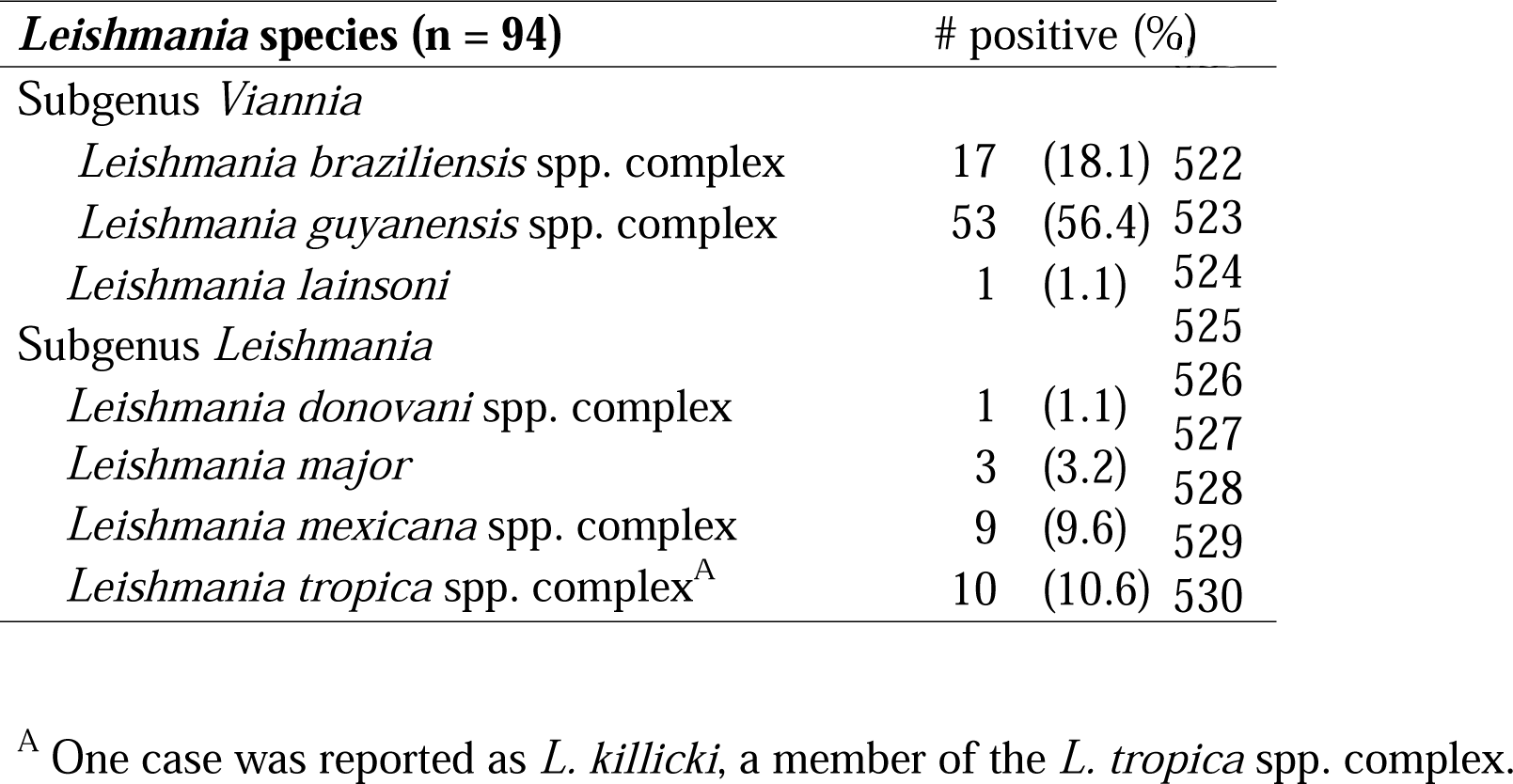
Species identified by mini-exon sequencing

### Demographics of positive cases

Of patients testing positive for *Leishmania*, 59/94 (62.8%) were aged 20-59 and 19 (20.2%) were under the age of 18, including 11 under age 10 years (Table 1, Figure S2). Sixteen patients (17.0%) were over age 60, including 1 patient over 80 years (Table 1, Figure S2). More patients with leishmaniasis were male (63.8%) (Table 1). Travel history was unavailable for 60 positive cases (63.8%), while 17 (18.1%) had documented international travel to *Leishmania*-endemic regions within 6 months prior to testing. Two (2.1%) cases documented international travel ≥5 years prior to diagnosis, while 7 (7.4%) noted travel history without specifying timing. Twelve positive cases (12.8%) were associated with travel to Costa Rica (Table S3). There were 7 cases (7.4%) with a history of migration from endemic areas and 1 case (1.1%) from a military service member with *L. major*. Although we did not receive cases where the possibility of endemic infection was explicitly documented, we noted 2 cases positive for *L. mexicana* spp. complex from Texas, where that species is endemic, although travel information was not provided.

### Specimen characteristics

Despite expectations for DNA degradation, formalin-fixed paraffin-embedded (FFPE) specimens yielded a higher overall positivity rate than fresh tissue specimens: 60.7% of 107 specimens versus 35.1% of 111 specimens (P < 0.0002, Chi-square test; Figure 2B, Table 3) (26). Positivity rate was highest (100%) in cases where staining patterns suspicious for intracellular organisms were reported (Table 3). Cases where characteristic *Leishmania* amastigotes were noted on histopathology had the second highest positivity rate (88.2%) (Table 3). Of the 2 cases that were histopathology positive but negative by molecular testing, one was instead positive for *Staphylococcus aureus* and the other for *Histoplasma capsulatum* by additional clinical molecular testing performed in our laboratory.

**Table 3.** Clinical and histopathological correlation with test positivity

|  | Positive for *Leishmania* (%) |
| --- | --- |
| **Specimen type (n = 218)** |  |
| Fresh tissue | 39/111 (35.1) |
| FFPE | 65/107 (60.7) |
| Total | 104/218 (47.7) |
| **Cases with ulcers or lesions on physical exam (n = 186)** |  |
| Yes | 45/68 (66.1) |
| Unknown | 49/118 (41.5) |
| **Cases with histopathological findings** |  |
| Amastigotes reported | 15/17 (88.2) |
| Staining suspicious for organisms | 12/12 (100.0) |
| No organisms seen | 27/61 (34.4) |
| Unknown | 40/96 (41.7) |

The most commonly involved body sites were skin, representing 86 cases (91.5%), particularly from exposed areas: arms, legs, head, and neck (Figure 3A). Four positive cases (4.3%) were detected from 5 oropharyngeal specimens and identified as *L. braziliensis* spp. complex, two of which were recurrent infections (Table S4). Only one specimen collected from internal organ sites or body fluids potentially associated with visceral disease tested positive: a paraffin-embedded inguinal lymph node positive for *Leishmania donovani* spp. complex (Figure 3B and 3C).

**Figure 3.**
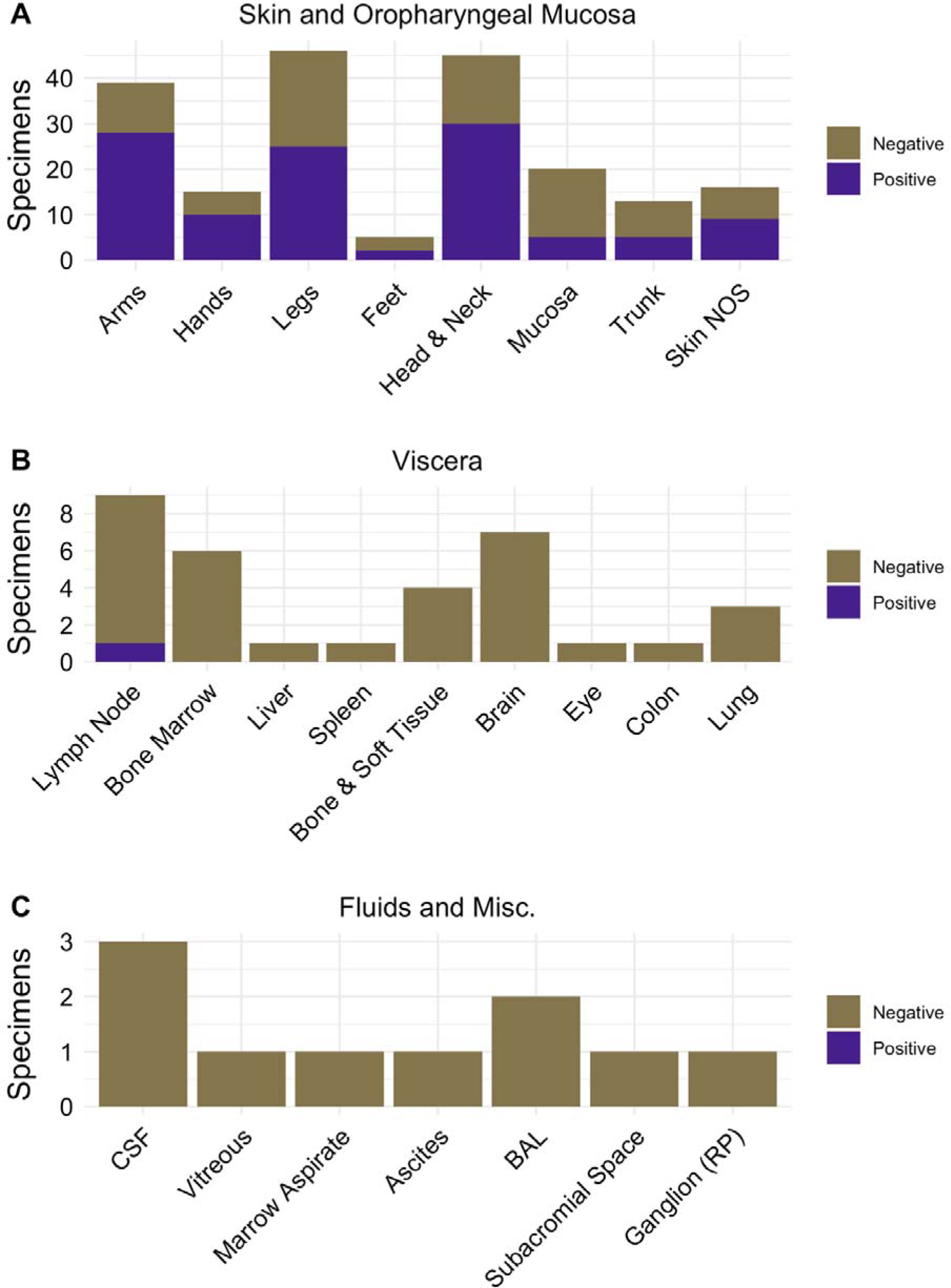
Specimen types submitted for *Leishmania* testing. Number of negative and positive specimens submitted from (A) skin and oropharyngeal mucosa, (B) internal organs, and (C) body fluids and miscellaneous sites. NOS = Not otherwise specified, RP = retroperitoneal.

22 patients had multiple specimens submitted. These included 13 positive cases and 9 negative cases (Table S4). Of the positive cases, 9 comprised specimen pairs collected less than 30 days apart. In 3 cases the initial specimen was positive, but subsequent specimens sent >30 days later were negative. One case involved multiple specimens collected 5 months apart that were positive for *L. guyanensis* spp. complex, having the same mini-exon sequence (Table S4). Two instances showed mini-exon sequence variation across different body sites: one case of *L. braziliensis* spp. complex in both an epiglottis and pharynx lesion and one case of *L. guyanensis* spp. complex in both an ear (helix) and shoulder lesion.

### Leishmania species and molecular epidemiology

Mini-exon sequencing provided adequate resolution (>80% bootstrap) to achieve species complex-level identification (Figure 4, Table 2). Of the 94 positive cases, 71 (75.5%) were subgenus *Viannia*. 53 (56.4%) were identified as *L*. *guyanensis* spp. complex, 17 (18.1%) as *L. braziliensis* spp. complex, and 1 (1.1%) as *L. lainsoni* spp. complex. Detected sequences included 18 previously unreported mini-exon alleles (Table S5, BioProject 974035).

**Figure 4.**
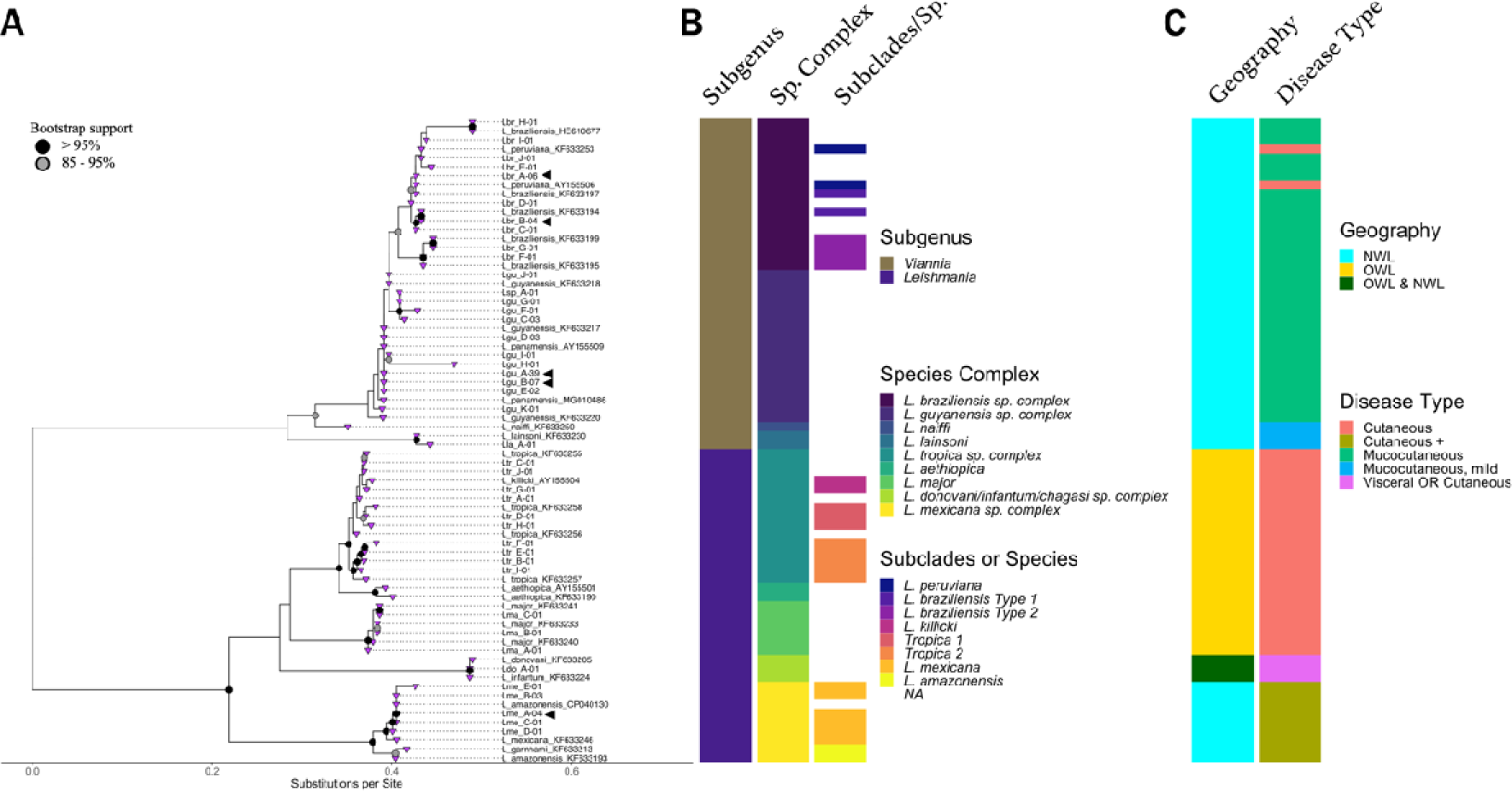
Taxonomic and epidemiologic characteristics of *Leishmania* mini-exon sequences. (A) Detected mini-exon sequences are represented as L, first two letters of species name, sequence variant, and number of times detected. The five most abundant mini-exon alleles are marked with arrowheads. Tips are purple triangles. Bootstrap values >95% are indicated as solid black nodes and 85-95% as grey nodes with black borders. Reference sequences are displayed with full species name and Genbank accession for mini-exon locus; *L. amazonensis* strain UA301 is indicated by the corresponding mini-exon locus CP040130. X-axis scale is substitutions per site. (B) Bars represent defined taxonomic descriptions of subgenus and species complex, as well as possible subclades or specific species within each complex as determined by mini-exon sequence similarity. (C) Epidemiologic associations, including geography and clinical presentation. OWL = Old World *Leishmania*, NWL = New World *Leishmania*.

We observed significant mini-exon sequence conservation within *L. braziliensis*. Two *L braziliensis* spp. complex alleles accounted for 10 of 17 cases (Lbr_A-06, Lbr_B-04) and were separated from each other by 3 polymorphisms. Sequence conservation across *L. braziliensis* and *L. peruviana* reference strains precluded resolution of these two species with two exceptions. Two mini-exon sequences, Lbr_F and Lbr_G, formed a clade (>95% bootstrap) with “Type 2” *L. braziliensis* mini-exon reference sequences (KF633199, MHOM/PE/03/LH2511; KF633195, MHOM/CO/90/LEM2216). Travel history was not available for Lbr_F, while Lbr_G had traveled to Peru. These Type 2 *braziliensis* sequences were detected once each.

The two member species of the *L. guyanensis* spp. complex, *L. guyanensis* and *L. panamensis*, had nearly identical previously published mini-exon sequences, precluding identification to the species rank. A single mini-exon allele (Lgu_A-39) accounted for most *L. guyanensis* spp. complex cases (33/53), including 6 cases with identical sequences recovered in multiple lesions. The second most abundant allele (Lgu-B-07) accounted for 7 cases and was only distinguished from Lgu_A-39 by homopolymer length variation. The next two most abundant alleles (Lgu_C-03, Lgu_D-03) accounted for 6 additional cases.

Nine cases (9.6%) were *L. mexicana* spp. complex and two alleles accounted for 7 of these (Lme_A-04 and Lme_B-03). The detected sequences formed a high-confidence clade with *L. mexicana* reference sequences (Figure 4). However, the mini-exon sequence (CP040130) from one recently completed, unpublished genome of *L. amazonensis*, UA301 (GCA_005317125.1) had 100% identity to multiple detected sequences. The UA301 mini-exon sequence thus defies the otherwise clear separation of *L. mexicana* and *L. amazonensis/L. garnhami* into two clades that are well-resolved by mini-exon sequences (Figure 4).

Fewer cases corresponded to OWL: 10 (10.6%) cases of *L. tropica* spp. complex and 3 (3.2%) *L. major*, one of which was from a military service member. Other patients positive for OWL include one patient who traveled to Yemen, one who had previously lived in and recently visited Tunisia, and one identified as an Afghan refugee (Table S3). Available clinical history for the one *L. donovani* spp. complex infection (1.1%) was not sufficient to determine its acquisition and molecular diagnosis could not resolve the organism to the species level. Thus, this case could represent either an infection with Old World species *L. donovani* or *L. infantum*, or infection with New World *L. chagasi/infantum*.

Sequences within the *L. tropica* spp. complex included two potential subclades with >80% bootstrap support (Figure 4A-B). The remaining allele (Ltr_G-01) had 99.75% identity over 392 nucleotides to *L. killicki* (GenBank AY155504) and was derived from a patient who had lived in and recently visited Tunisia. This case was reported to the species rank given the geographic association with *L. killicki* (Figure 4A, Table S3) (27).

The distribution of identified species paralleled the travel history in the 34 cases with documented exposures. Of these, 23 (67.6%) had traveled to or from North, Central, or South America (Table 2, Table S3). There were 7 known migration-related cases, including 6 NWL infections, comprising five *L. guyanensis* spp. complex; 1 *L. braziliensis* spp. complex. Four involved patients had traveled from Central and/or South America, while one had immigrated from West Africa, but did not have explicitly documented travel history to an endemic area. One patient from Afghanistan tested positive for *L. tropica* spp. complex (Table S3, Table S5).

## Discussion

Our work describes molecular reference testing for *Leishmania* in the general U.S. population during a period where it was otherwise unavailable. This has provided a unique opportunity to compile a comprehensive, 1-year catalog of patient and pathogen characteristics that help approximate the incidence and epidemiology of infections in the U.S. In agreement with prior studies involving U.S. travelers, our case demographics reflect a slight male predominance at 63.8% (12,28–30). Also consistent with previous observations, positive cases were associated with recent travel to Central and South America, particularly Costa Rica, where there is a high incidence of leishmaniasis (Table 1, Table S2) (12,28,29,31). 11.7% of cases in this work were in children younger than 10 and 6.4% in the 10-18 age range, adding to reports of pediatric infection in the U.S. (12,13,29,32,33)

Migration-related leishmaniasis has increased globally, but U.S.-specific data has been limited to sporadic case studies (28,34–37). This work identifies 7 cases (7.4% of all positive) of likely migration-related infection originating from multiple continents, highlighting a need for increased clinician awareness in this vulnerable population (Table 1, Table S3). Given that travel histories were not available for most (63.8%) cases, this likely underestimates the true disease burden in this group. Endemic infection with *L. mexicana* spp. complex has been documented in Texas and Oklahoma, but the provided clinical histories from our cases did not enable us to determine the probability of autochthonous leishmaniasis.

CL is the most common clinical manifestation reported in recent U.S. studies. (11–13), and 91.5% of cases in this study were CL. We additionally detected *L. braziliensis* in 4 patients with mucosal disease, including two with relapsed MCL acquired during travel several years prior. In contrast, a prior case series and review by Murray et al. reported only 6 documented civilian cases of MCL in the U.S from 1975-2020 (36). Our cohort may be enriched for MCL due to the high prevalence of subgenus *Viannia*, which accounted for 75% of all cases.

Identification to species complexes within subgenus *Viannia* is clinically informative for treatment and monitoring, due to the associated risk of progression to MCL with most *Viannia* species (3). Within *Viannia*, mini-exon sequencing typically lacked resolution beyond the species complex level, particularly *L. guyanensis* spp. complex (Figure 4). Consistent with prior analyses (6), our assay could distinguish *L. braziliensis* Type 2 from *L. peruviana* (Figure 4). Most Type 1 *L. braziliensis* and *L. peruviana* couldn’t be distinguished; clinically, this may overestimate a patient’s risk of developing mucosal disease.

Intriguingly, we identified a high-confidence clade of *L. braziliensis* Type 1 (KF633199, Lbr-B, Lbr-C) that may be separable from *L. peruviana* and an outlier sequence in *L. braziliensis* spp. complex (Lbr_H) with 100% identity to the first reported *L. braziliensis* infection in Suriname (HE610677, Figure 4) (38). These emerging clades suggest additional mini-exon sequences from well-characterized strains could increase molecular diagnostic resolution, particularly in combination with detailed travel history. We note several patients with positive cutaneous specimens who traveled to multiple countries and were consequently at risk for infection from different *Leishmania* species. In such instances improved species-level identification would more accurately assess risk for progression to mucosal involvement (3).

Visceral leishmaniasis cases are rare in the U.S. but have been documented in service members returning from endemic regions, and there are concerns for undetected asymptomatic infection and risk for reactivation in these veterans (11,12,39). In this series, only one case tested positive for *L. donovani* spp. complex, which is associated with visceral disease and was consistent with our detection in a lymph node. In agreement with previous studies (6) our assay could not distinguish between *L. donovani* and *L. infantum/chagasi*.

Clinical and laboratory diagnosis of leishmaniasis can be challenging, and molecular methods are considered the most sensitive diagnostic modality (2,3). This work and others have demonstrated the utility of the mini-exon gene as a sensitive, specific, and effective target for distinguishing the major species complexes (6,7,40). Moreover, our assay readily detects infection from a variety of specimen types associated with CL, MCL, and VL, and has demonstrated good performance with FFPE specimens. With rapid turnaround times (∼3 days), molecular testing and sequencing can effectively inform patient treatment and management.

We found that mini-exon sequencing is sufficient for identification of clinically relevant species complexes, supporting prior work showing that species identification from mini-exon analysis had comparable performance to multilocus sequence typing or single gene target assays (6,40). We successfully resolved *L. lainsoni* from other *Viannia* species, which has historically posed technical challenges (6). However, an important limitation is that most closely related species within each of the *L. donovani, L. guyanensis,* and *L. braziliensis* species complexes could not be resolved. This challenge is clinically relevant within *L. braziliensis* spp. complex, given the greater propensity of *L. braziliensis* for mucocutaneous spread relative to *L. peruviana* (41). Nevertheless, expanding publicly available mini-exon sequence records may enable more specific identification.

We identified 18 previously unreported mini-exon alleles, expanding the diversity of known sequences and highlighting opportunities to improve molecular species identification. Whole-genome sequencing will enable more accurate and specific identification, resolve technical artifacts, and identify co-infections (7). The challenges in resolving species within complexes and unexpected findings from recently sequenced strains, such as *L. amazonensis* UA301 clading with *L. mexicana* strains, emphasize the evolutionary complexity of *Leishmania* species (42–44).

Although illuminating, there are several limitations to this study. First, our estimates of disease burden and the granularity of our demographic data are limited by the incompleteness of clinical information provided to our reference laboratory. This poses particular challenges in interpreting patients with multiple positive specimens, as prolonged PCR positivity does not necessarily reflect treatment failure (45). Second, it is likely that additional civilian cases were not tested at our laboratory, particularly after testing resumed at CDC during July 2022. Third, *Leishmania* testing remained available for Department of Defense personnel and contractors at the Walter Reed Army Institute of Research (WRAIR) during the study period (46); therefore, the single *L. major* case we detected in a U.S. armed forces personnel likely underrepresents disease burden. A survey by Stahlman *et al* documented over 2,000 cases in the U.S. Armed Forces from 2001-2016 but noted significantly fewer cases starting in 2011, with only 11 in 2015 (3,11,46). Finally, mini-exon sequence data have inherent limitations. The *Leishmania* mini-exon gene is comprised of 100-200 tandem repeats, which increases the risk of chimeric PCR artifacts (7) and masks low abundance sequence variants. Although our validation studies confirmed this issue did not interfere with the ability to discriminate between different species complexes, we are limited in making more refined phylogenetic inferences that would require more comprehensive whole-genome sequencing data.

Leishmaniasis remains challenging to recognize clinically, and we expect it is still underdiagnosed in the U.S. (2,12,13). Most state public health agencies do not require positive test notification, with the exception of Texas, and suboptimal reporting compliance has been documented (13). Our study demonstrates that molecular testing by mini-exon sequencing provides clinically actionable species-level identification, and when considered in aggregate, suggests a significantly higher annual incidence in the U.S. than previously appreciated. The burden, distribution, and biology of this disease will continue to change with global travel, migration, and climate changes impacting parasite, vector, and host geography (47). This work highlights the role of molecular testing in enabling diagnostic laboratory professionals, providers, and public health agencies to track and appropriately treat this disease.

## Acknowledgements

We are grateful to Stephanie Krieg, MLS(ASCP) and Dr. Dhruba SenGupta for their work establishing clinical assay in the laboratory information system (LIS) and integration with the electronic medical record; Dan Hoogestraat MB(ASCP) for assistance obtaining data from the LIS; and Dr. Noah Hoffman for helpful discussions of phylogenetic analysis. We thank our colleagues at the CDC Parasitic Disease Branch for referring testing to the University of Washington.

## Biographical Sketch

Dr. Truong is a clinical microbiology and molecular microbiology laboratory director at UW Medicine. Her research interests include diagnostic stewardship and evaluating the clinical impact of rapid diagnostic technologies to improve patient care.

## Supplemental Materials

**Figure S1.**
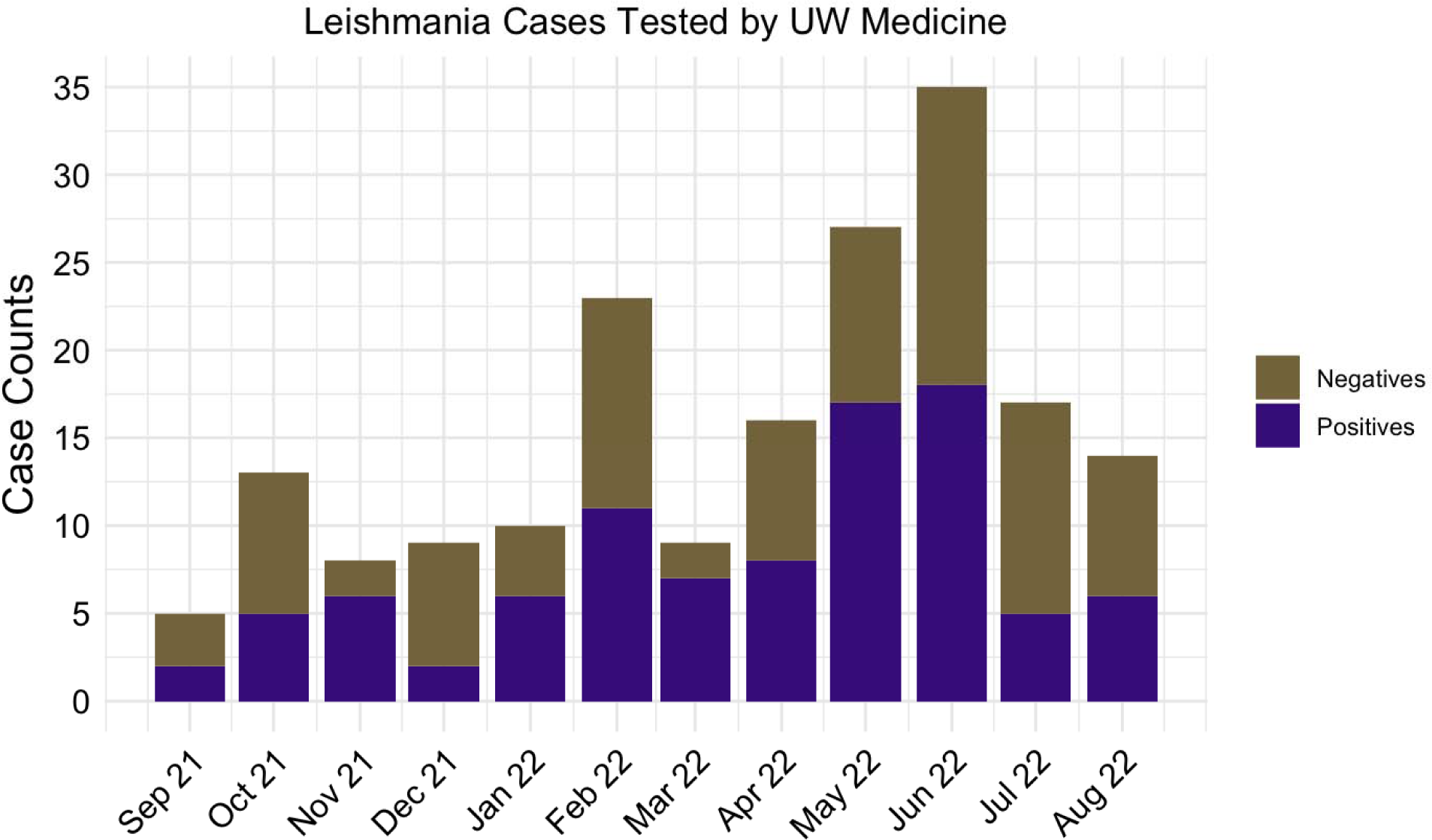
Case volume and positivity by month of testing. Number of negative and positive specimens submitted and tested during each month of the study period. Negative cases are in gold; positive cases in purple.

**Figure S2.**
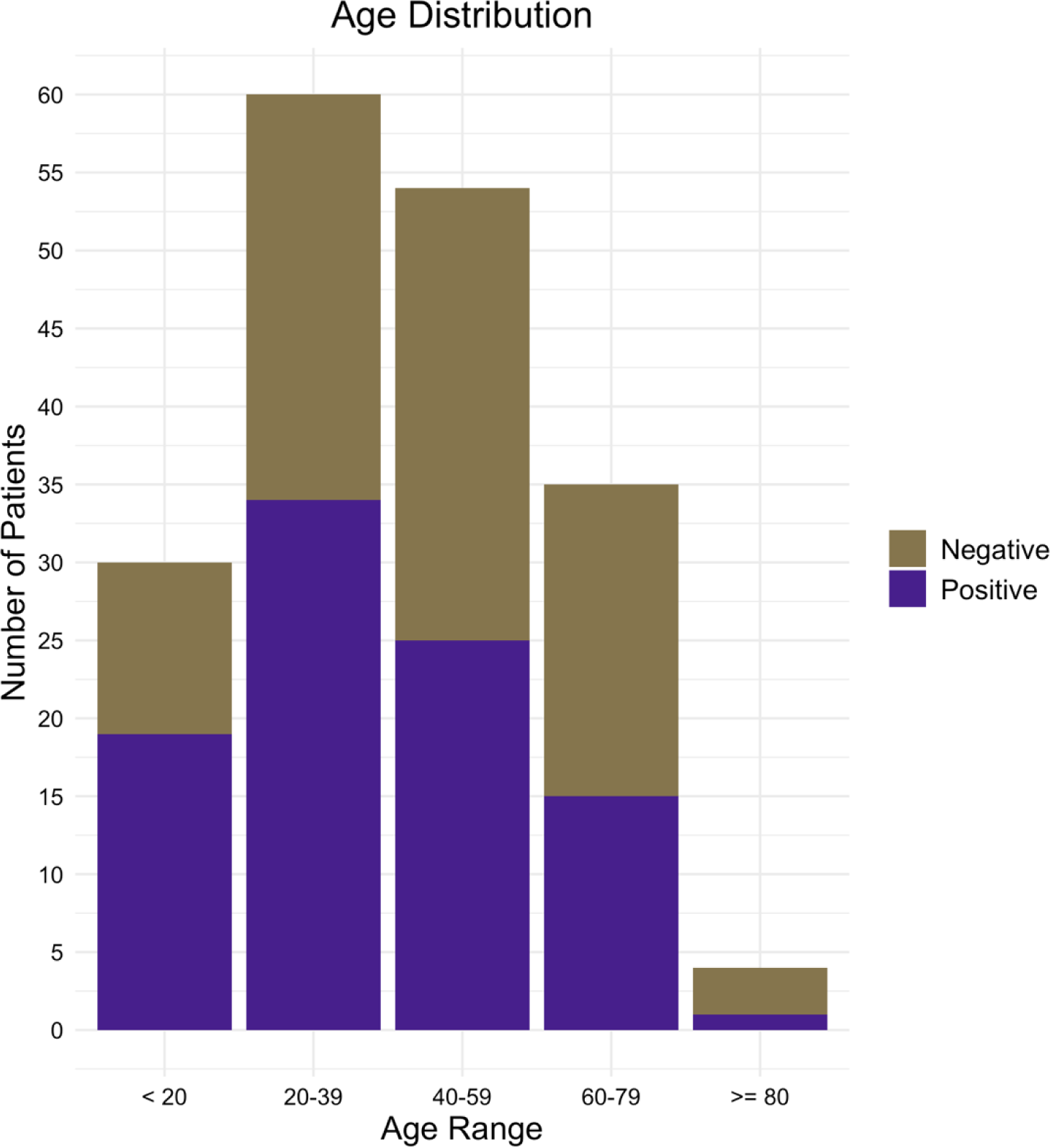
Age Distribution of Patients Tested for Leishmaniasis. Number of patients tested by indicated age range (X-axis). Negative cases are in gold; positive cases in purple.

**Table S1.** Test performance and validation

| Specimen number | Specimen type | Reference identification | *Leishmania* PCR identification | Agreement |
| --- | --- | --- | --- | --- |
| 1 | Cultured strain | *L. donovani* | *L. donovani* spp. complex | Yes |
| 2 | Cultured strain | *L. major* | *L. major* | Yes |
| 3 | Cultured strain | *L. aethiopica* | *L. aethiopica* | Yes |
| 4 | Cultured strain | *L. chagasi* | *L. donovani* spp. complex | Yes |
| 5 | Cultured strain | *L. guyanensis* | *L. guyanensis* | Yes |
| 6 | Genomic DNA | *L. tropica* | *L. tropica* | Yes |
| 7 | Cultured strain | *L. panamensis* | *L. guyanensis*/*L. panamensis* | Yes |
| 8 | Cultured strain | *L. amazonensis* | *L. amazonensis* | Yes |
| 9 | Patient specimen FFPE | *L. tropica* | *L. tropica* | Yes |
| 10 | Patient specimen FFPE | *L. braziliensis* | *L. braziliensis* | Yes |
| 11 | Patient specimen FFPE | *L. braziliensis* | *L. braziliensis* | Yes |
| 12 | Patient specimen FFPE | *L. braziliensis* | *L. braziliensis* | Yes |
| 13 | Patient specimen FFPE | *L. braziliensis* | Not detected | No |
| 14 | Patient specimen FFPE | *L. braziliensis* | *L. braziliensis* | Yes |
| 15 | Patient specimen FFPE | *L. braziliensis* | *L. braziliensis* | Yes |
| 16 | Patient specimen FFPE | *L. braziliensis* | Not detected | No |
| 17 | Patient specimen FFPE | *Leishmania* sp. | *L. chagasi*/*L. infantum* | Yes |
| 18 | Patient specimen tissue | *Leishmania* sp. | *L. guyanensis*/*L. panamensis* | Yes |
| 19 | Patient specimen tissue | *Leishmania* sp. | *L. guyanensis*/*L. panamensis* | Yes |
| 20 | Patient specimen FFPE | *Leishmania* sp. | *L. guyanensis*/*L. panamensis* | Yes |
| 21 | Patient specimen tissue | *Leishmania* sp. | *L. braziliensis*/*L. peruviana* | Yes |
| 22 | Patient specimen fluid | *Pneumocystis jiroveci* | Not detected | Yes |
| 23 | Patient specimen fluid | *Pneumocystis jiroveci* | Not detected | Yes |
| 24 | Patient specimen tissue | *Toxoplasma gondii* | Not detected | Yes |
| 25 | Patient specimen tissue | *Toxoplasma gondii* | Not detected | Yes |
| 26 | Patient specimen tissue | *Histoplasma capsulatum* | Not detected | Yes |
| 27 | Patient specimen tissue | *Histoplasma capsulatum* | Not detected | Yes |
| 28 | Patient specimen FFPE | *Histoplasma capsulatum* | Not detected | Yes |
| 29 | Patient specimen fluid | *Histoplasma capsulatum* | Not detected | Yes |
| 30 | Patient specimen fluid | *Candida albicans* | Not detected | Yes |
| 31 | Patient specimen FFPE | *Candida tropicalis* | Not detected | Yes |
| 32 | Patient specimen fluid | *Candida glabrata* | Not detected | Yes |
| 33 | Patient specimen tissue | *Sporothrix schenckii* | Not detected | Yes |
| 34 | Patient specimen tissue | *Aspergillus fumigatus* | Not detected | Yes |
| 35 | Patient specimen FFPE | *Aspergillus fumigatus* | Not detected | Yes |
| 36 | Patient specimen tissue | *Escherichia coli* | Not detected | Yes |
| 37 | Patient specimen FFPE | *Escherichia coli* | Not detected | Yes |
| 38 | Patient specimen tissue | *Pseudomonas aeruginosa* | Not detected | Yes |
| 39 | Patient specimen tissue | *Pseudomonas aeruginosa* | Not detected | Yes |
| 40 | Patient specimen fluid | *Staphylococcus aureus* | Not detected | Yes |
| 41 | Patient specimen fluid | *Staphylococcus aureus* | Not detected | Yes |
| 42 | Patient specimen tissue | *Streptococcus pneumoniae* | Not detected | Yes |
| 43 | Patient specimen fluid | *Streptococcus pneumoniae* | Not detected | Yes |

**Table S2.**
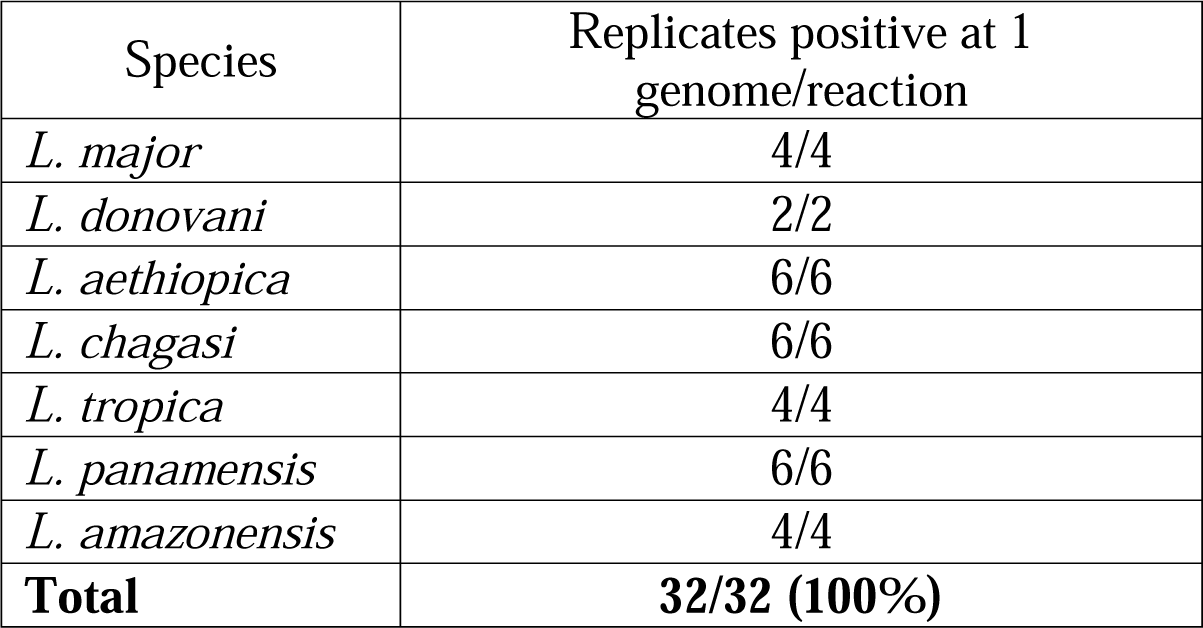
Limit of detection

**Table S3.**
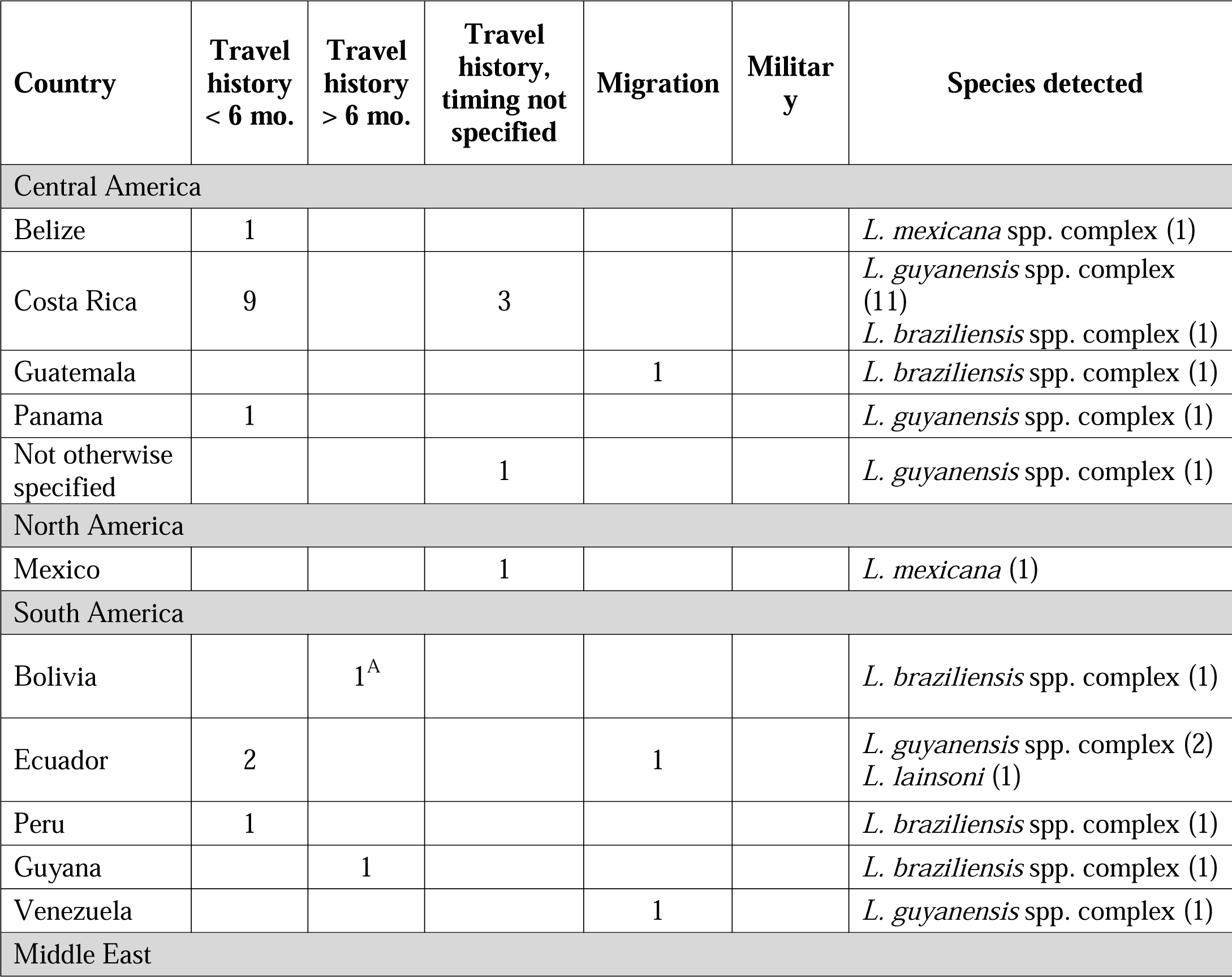
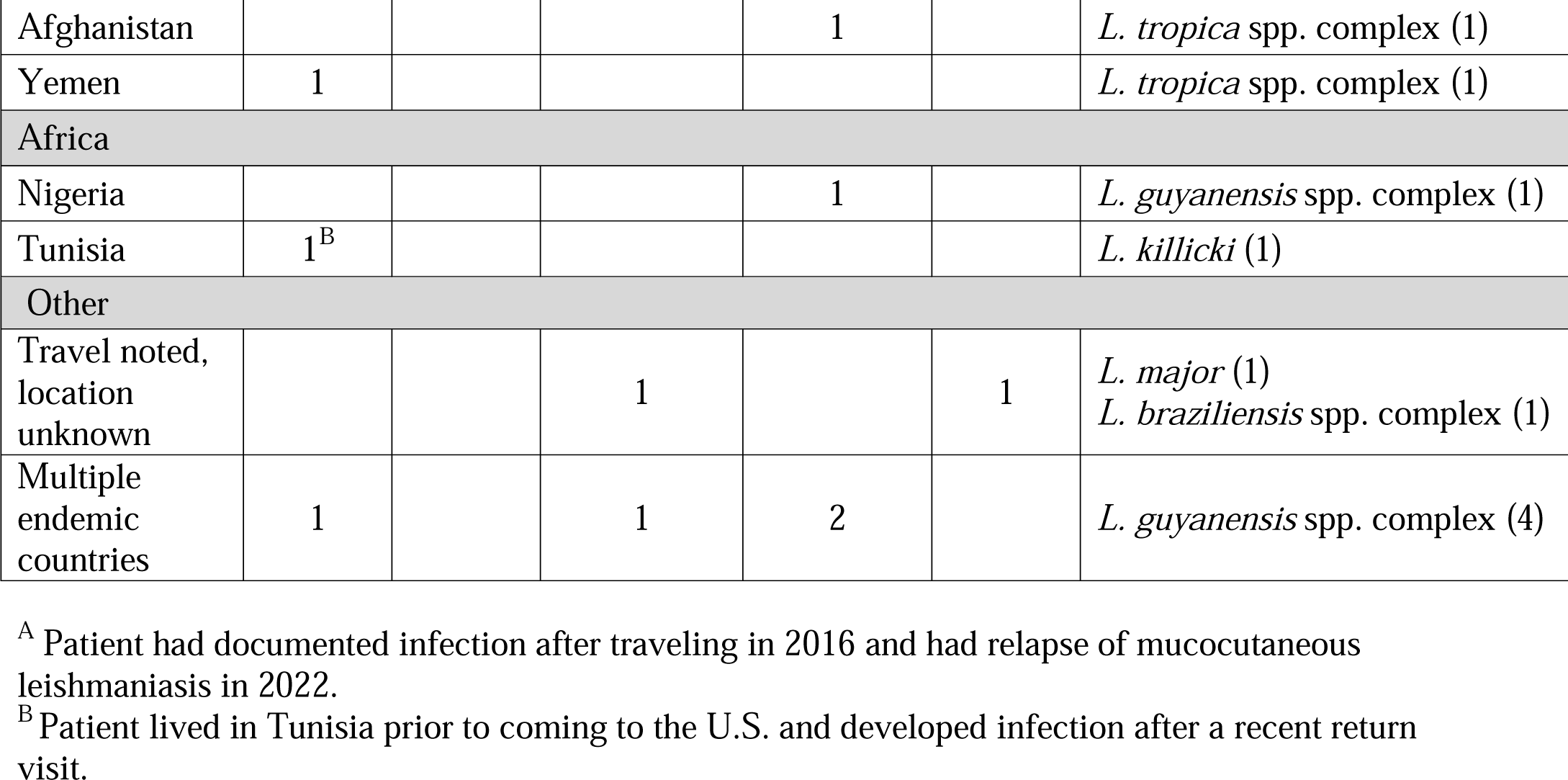
Species identified by travel history

**Table S4.** Repeat testing

| Category | # patients |
| --- | --- |
| I. Multiple specimens negative |  |
| a. 30+ days apart | 3 |
| b. Within 30 days | 6 |
| II. Multiple specimens positive within 30 days |  |
| a. Different sites | 4 |
| b. Similar or same site | 4 |
| c. Unknown if sites are same or different | 1 |
| III. Initial positive specimen, subsequent negative specimen 30+ days later | 3 |
| IV. Multiple specimens positive 30+ days apart | 1 |

**Table S5:**
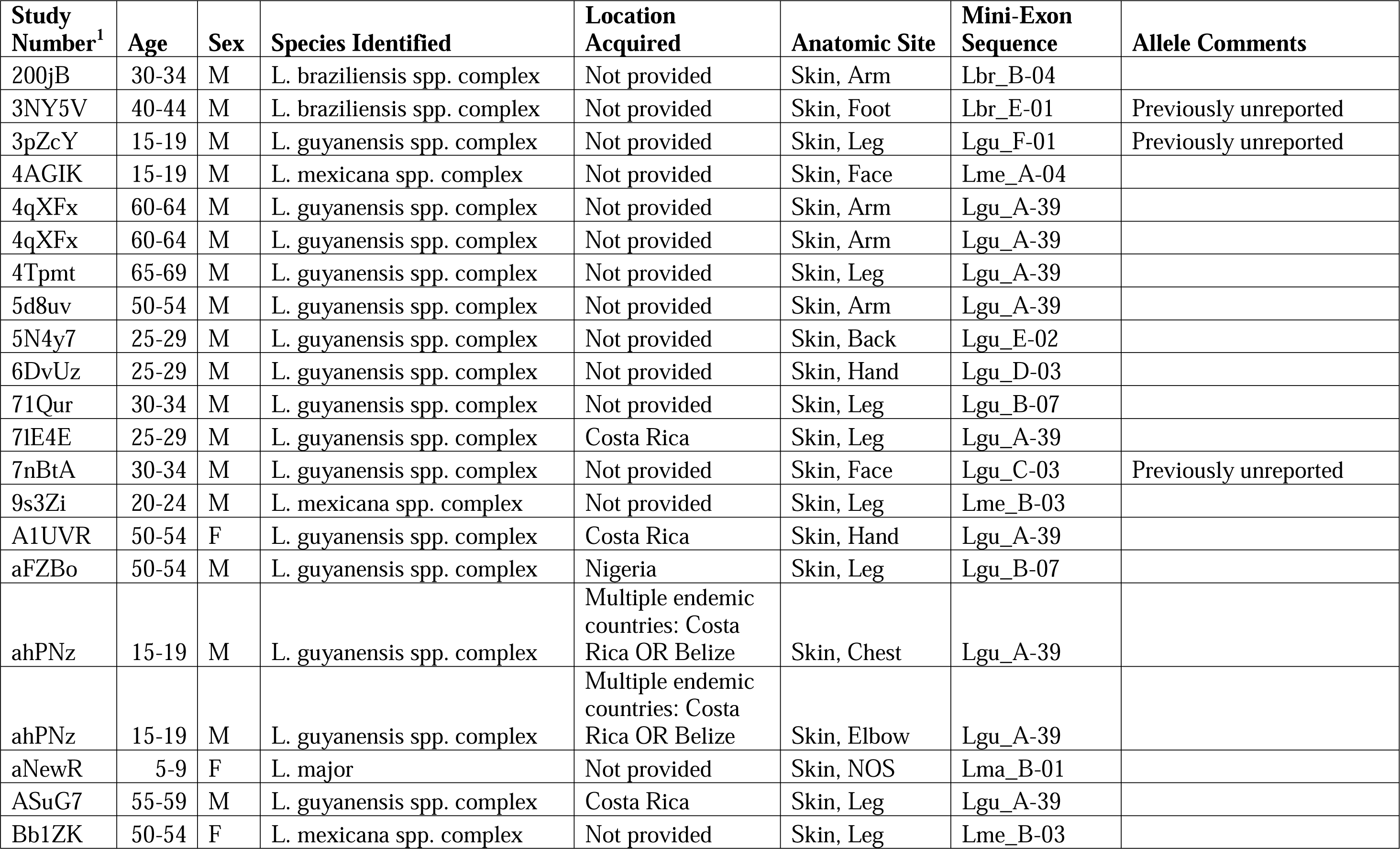
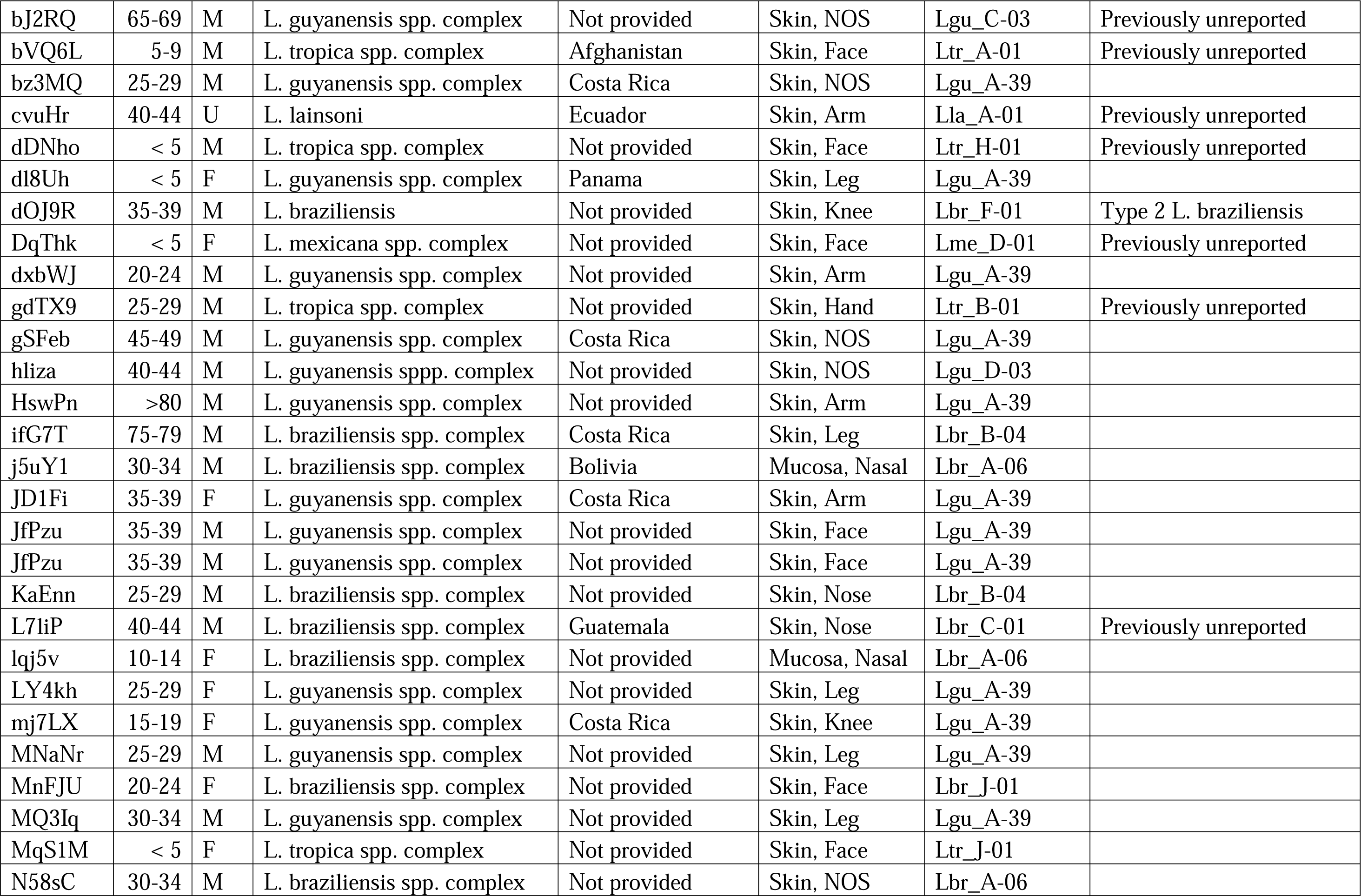
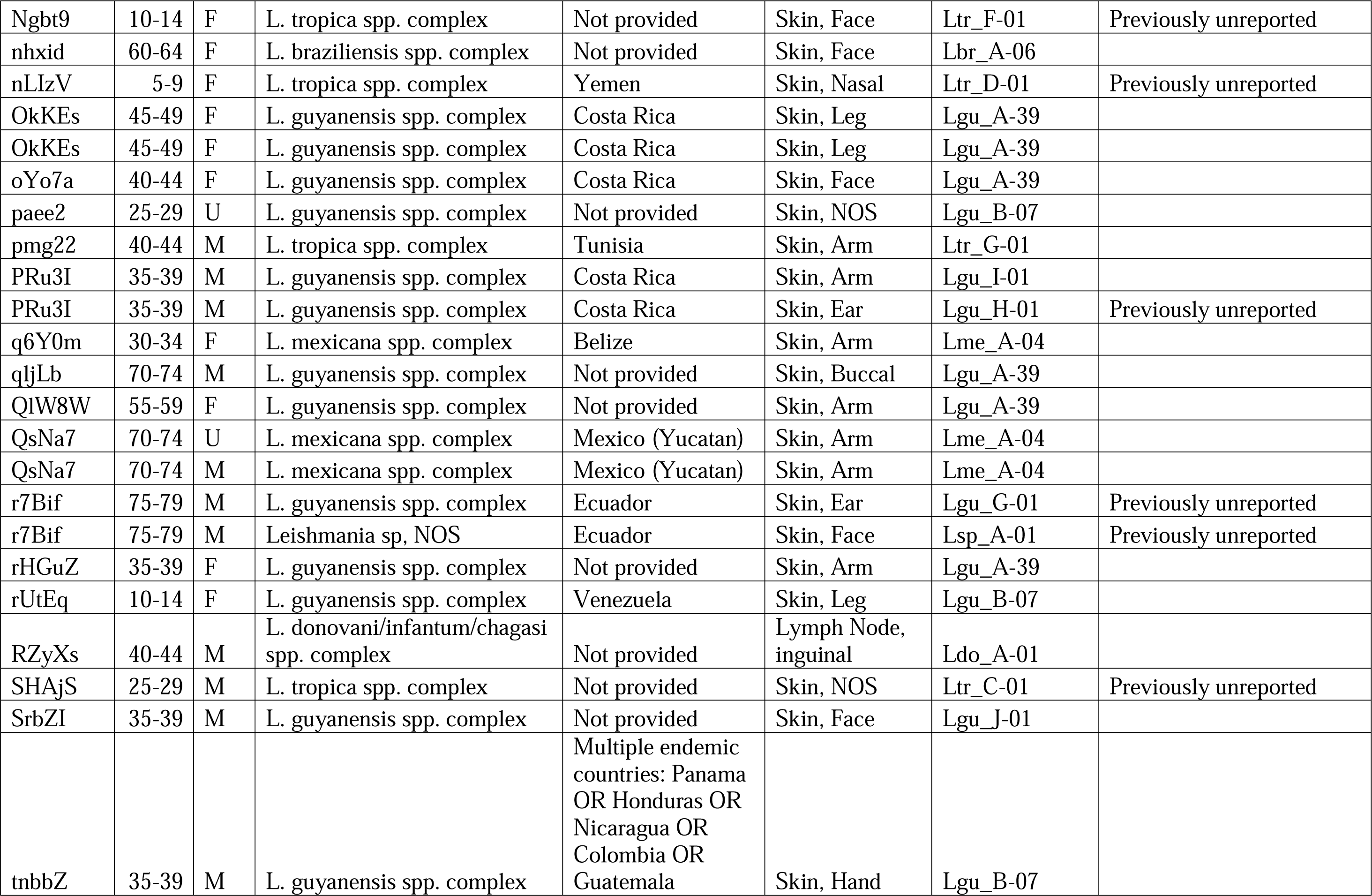
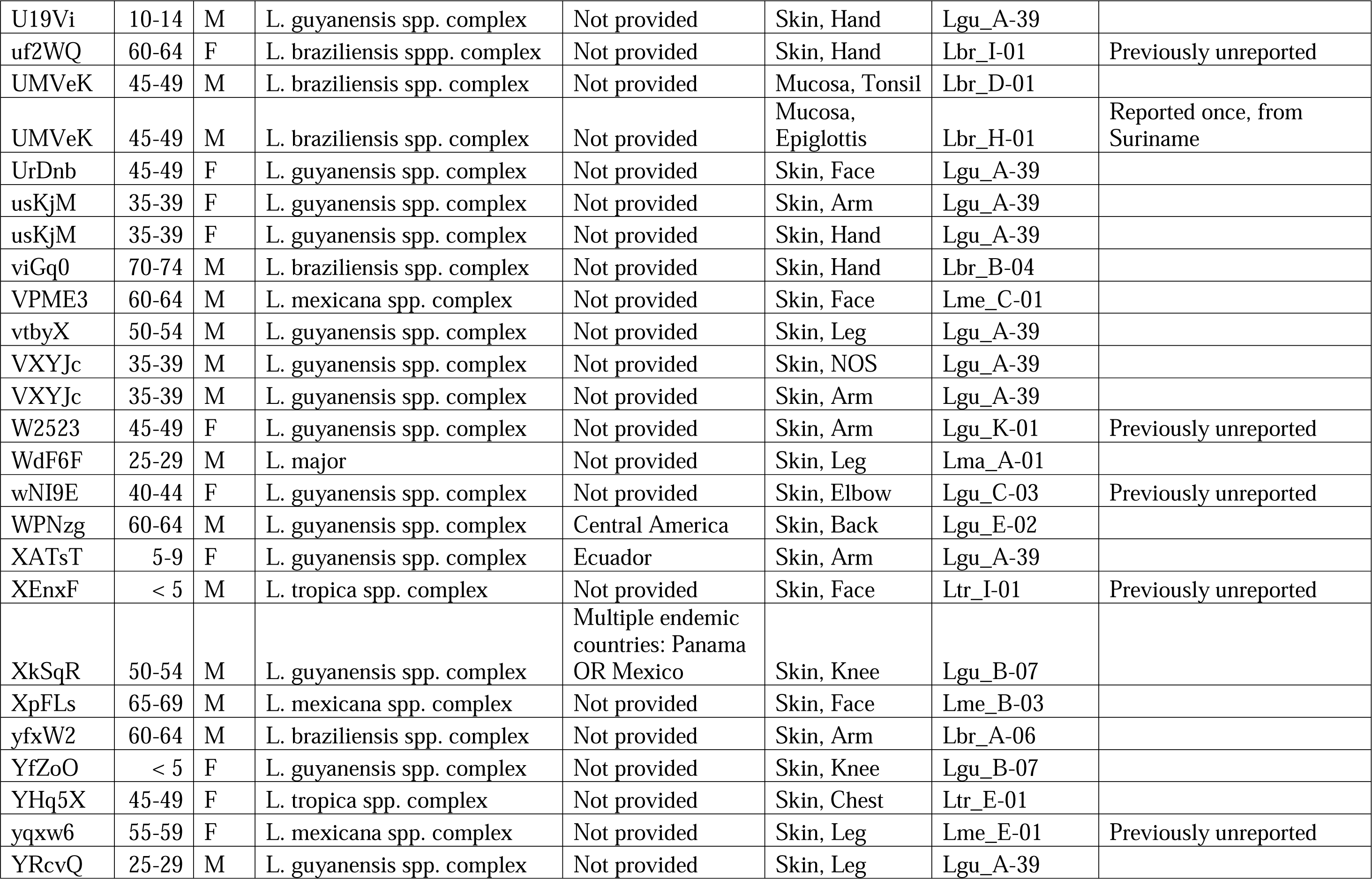
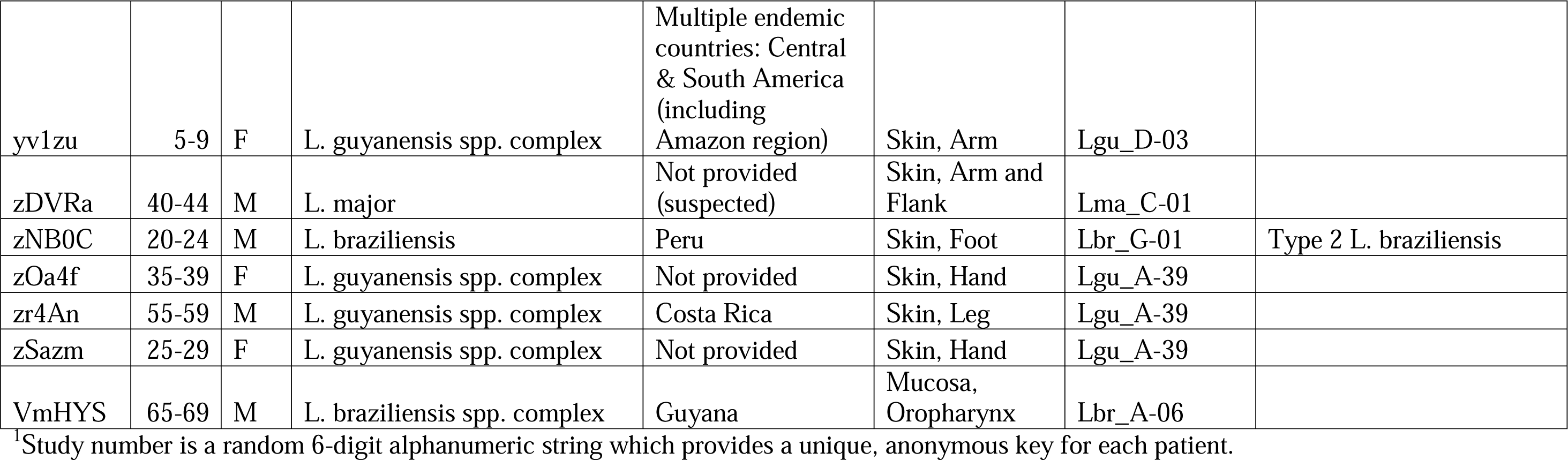
Metadata for Positive Cases

